# Climate and topography shape malaria transmission in lowland Busia and highland Meru counties, Kenya

**DOI:** 10.64898/2026.06.30.26356813

**Authors:** Bryson Alberto Ndenga, Gladys Adhiambo Agola, Kevin Omondi Owuor, Joel Omari Mbakaya, Charles Ochieng Ronga, Edith Chepkorir, Lydiah W. Kibe, Hoseah Miima Akala

## Abstract

**Introduction:** Malaria is a global public health problem especially in sub-Saharan Africa. In Kenya, it varies across ecological zones with limited evidence comparing vector ecology, climate, topography and risk of infection in western lowlands and central highlands.

**Objective:** To compare levels of malaria infection, vector densities, rainfall, temperature, relative humidity and topography in lowland Busia and highland Meru counties.

**Methods:** A cross-sectional survey was conducted using larval dipping for aquatic habitats, pyrethrum spray catches for indoor resting mosquitoes, and malaria diagnosis using rapid diagnostic tests and microscopy. Rainfall, temperature, and relative humidity were recorded using automated data loggers. Topography was noted by ground truthing. Data were analysed using chi-square tests, analysis of variance, and logistic regression.

**Results:** Early instar *Anopheles* larvae were significantly less likely to be detected in the highland site than in the lowland site (uOR = 0.33; 95% CI: 0.11–0.97). Malaria prevalence by rapid diagnostic tests was 0% in the highland site and significantly higher in lowland sites (p<0.001), with microscopy confirming the absence of infections in the highland area. Highland sites experienced significantly cooler temperatures, including more hours below 16°C (p=0.006), whereas lowland sites recorded significantly higher minimum, mean, and maximum temperatures (p<0.001). Rainfall did not differ significantly between the two ecological zones (p=0.090), average minimum RH in the highland was significantly higher than in the lowland site (p<0.001). Valleys in Baragu are mainly V-shaped while Maduwa and Budalangi are generally flat areas in the lower courses of rivers prone to flooding.

**Conclusion:** Cooler highland climates and topographic features likely limit vector presence, abundance, development and malaria transmission, while warmer lowland environments sustain residual transmission.

## Introduction

In the year 2024, the number of malaria cases globally was estimated at 282 million, an increase of about nine million cases compared with 2023, most of which were reported in Ethiopia, Madagascar and Yemen [1]. In the past decade, malaria cases have increased with most reports being from the WHO African Region. The estimated global deaths in 2024 were 610 000, with over a half of them being reported in four countries, namely, Nigeria, the Democratic Republic of the Congo, the Niger and the United Republic of Tanzania.

In Kenya, malaria is a significant cause of morbidity and mortality, an estimated six million cases are reported annually with 70% of the population at risk of infection [2]. In the year 2022, over four million malaria cases and 769 deaths were reported [3]. Most (79%) of these malaria cases occurred in the lake endemic area, followed by the coastal endemic (11%), then in highland epidemic (7%) and the least (3%) in seasonal zone [4]. In the year 2020, malaria prevalence among children under five years in Kenya was at 6% [3]. The risk of being infected with malaria in Kenya varies geographically as influenced by altitude, rainfall patterns, and temperature and is categorized into seven strata, namely: no transmission; very low burden; low burden; moderate burden; moderate to high burden; high burden and very high burden [5]. Increase in malaria incidence and mortality in some parts of Kenya is associated with increased rainfall, temperature and humidity [6,7]. However, in other parts of the country, an increase in air temperature negatively affects mosquito aquatic habitats and the life cycles of both malaria vectors and parasites [8].

Malaria is endemic in Busia County [9,10] with a hospital prevalence of 32-37% [11,12]. The most common malaria parasite within the western Kenya region, where Busia County is located, is *Plasmodium falciparum* although *P. malariae* and their co-infections do occur [12]. Meru County is located within the low risk malaria transmission central Kenya highlands where the disease is not reported in most areas [13,14].

Temperature, rainfall, and humidity are three main climatic parameters that determine mosquito survival [15]. An increase in temperature accelerates the rate at which mosquito larvae develop, the frequency at which adult females blood feed on humans, and reduces the time it takes for malaria parasites to develop and mature in female mosquitoes and become infective [16]. The annual mean minimum and maximum temperatures required for *P. falciparum* transmission is 16°C and 34°C [16–19]. The survival of *adult An. gambiae s.s.* is highest at 15-25°C and at 60-100% relative humidity [18].

Topography is an important physical feature that defines the risk of local malaria transmission in an area [20–22]. In the western Kenya highlands, *Anopheles* larval density, adult indoor resting densities, entomological inoculation rates, prevalence of asymptomatic malaria, immune response to malaria and the spatial distribution of malaria infections are higher in broad flat bottomed “U-Shaped” valleys than in narrow “V-Shaped” valleys during both the dry and rainy seasons [20–22]. In lowland plains, especially in the lower courses of rivers, floods that occur during the rainy seasons usually wash away mosquito aquatic stages, a situation known as the “flushing effect”, that reduces both malaria vectors and cases [23]. However, when flood waters subside forming pools, malaria vectors re-establish themselves and can lead to increased malaria transmission [24].

Despite evidence that climate and topography influence malaria transmission in Kenya [20–22,25,26], comparative data integrating entomological, epidemiological, and climatic measurements contrasting western lowlands with central highlands in Kenya remain limited. Therefore, this study aimed to determine and compare the recent levels of malaria infection, vector densities, rainfall, temperature and relative humidity in lowland Busia and highland Meru counties in Kenya.

## Methods

### Study design

This was a cross-sectional study conducted during the short rainy season and early dry season to capture early malaria transmission dynamics following seasonal rainfall.

### Study sites

Two study sites were identified on the Equator using a Global Positioning System (GPS) unit (Garmin eTrex® 10, Garmin International Inc. Kansas, USA) in Busia (Maduwa site) and Meru (Baragu site) counties in Kenya for field data collection activities (Fig 1). A sampling area measuring 1km x 1km was demarcated in these two sites. A third site (Budalangi and its surrounding villages) also in Busia County was selected to screen adult volunteers for malaria infection (Fig 1). Coordinate readings (latitude and longitude) and altitude of houses, schools, churches and other major structures were taken using the GPS unit.

**Fig 1.**
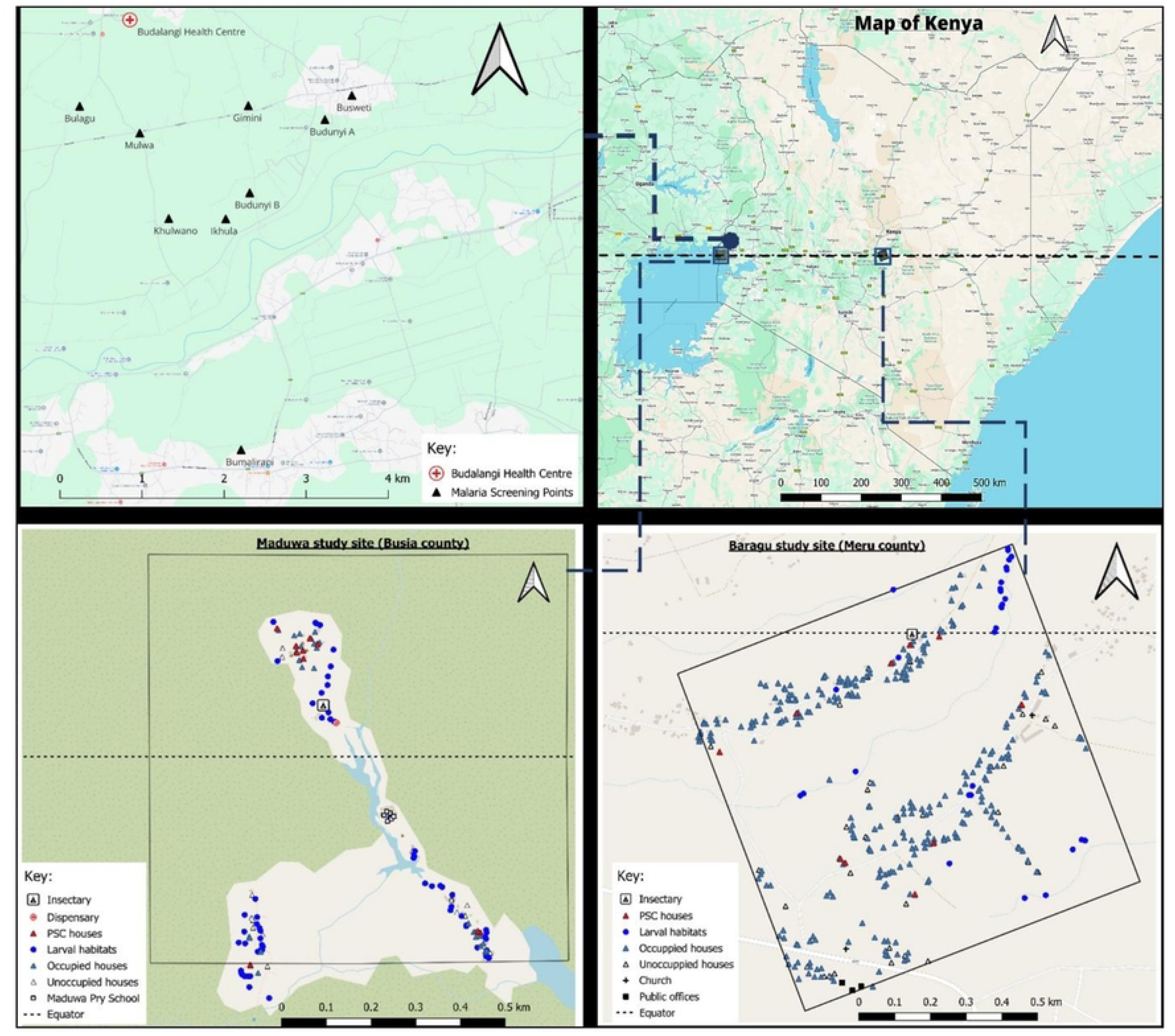
Study site map of Maduwa and Budalangi in lowland Busia County and Baragu in highland Meru County in Kenya.

Maduwa site (S00.00000°, E034.02051°; 1,123-1,142 metres above sea level is located within the Yala swamp close to Lake Victoria and near Osieko, Bunyala Sub-County. It is approximately 43.7 km south of Busia Town, the headquarters of Busia County. Fishing and small-scale agriculture are the main economic activities in this village. Crops grown include maize, beans, vegetables, cassavas, potatoes and bananas. Livestock kept are mainly indigenous cattle, pigs and poultry. This area is prone to flooding during both long and short rainy seasons when River Yala breaks its banks upstream. Several residential houses have been destroyed by flooding forcing some residents to relocate to other places mainly to Osieko Market. A local dispensary was closed due to flooding. Hence residents travel to Osieko Dispensary for their medical care. Classrooms, offices and teachers’ houses in Maduwa Primary School experience flooding during the rainy seasons. Means of transport are mainly by boats, either mechanical or engine driven.

Baragu site (N00.00000°, E037.67229°; 1,411-1,531 m) is located near Gatimbi Market in Meru Central Sub-County. It is approximately 6.8 km south of Meru Town, the headquarters of Meru County. Farming is the main economic activity. Coffee and bananas are the main cash crops. Other crops grown include maize, beans, vegetables, cassavas and potatoes. Livestock kept are mainly dairy cattle and poultry. Most valleys in Baragu are very steep (V-shaped) with numerous streams. Most residents go to Gatimbi Health Centre for medical services while a few others travel to Meru Town to public or private hospitals.

Budalangi site, which includes the Budalangi Health Centre is located 17.7 kilometres from the Maduwa site. The nine malaria screening sites for adult participants only included: Gimini (N00.12272°, E034.04366°; 1,127 m); Khulwano (N00.11027°, E034.03495°; 1,150 m); Ikhula (N00.11023°, E034.04119°; 1,146 m); Budunyi B (N00.11311°, E034.04382°; 1,143 m); Mulwa (N00.11970°, E034.03178°; 1,148 m); Bumalirani (N00.08478°, E034.04287°; 1,142 m); Bulagu (N00.12266°, E034.02520°; 1,145 m); Budunyi A (N00.12119°, E034.05205°; 1,173 m) and Busweti (N00.12385°, E034.05500°; 1,151 m). These sites are located in Bunyala Sub-County in Busia County. In all these villages in Budalangi, altitude ranged from 1,127-1,173 m. Most parts of Budalangi are generally flat and prone to flooding during the rainy seasons with several raised hilly places.

Ground truthing by physically walking within the selected sites in Maduwa, Baragu and Budalangi was done to ascertain the landscapes and shapes of valley bottoms and described them as either broad flat bottomed “U-Shaped” valleys, narrow “V-Shaped” valleys or flat plains.

### Larvae/pupae sampling

To determine the presence of mosquito larvae/pupae, sampling was done once using a standard 350 ml capacity mosquito dipper (Clarke Corporation, Illinois, USA). This was carried out on 9^th^ November 2024 in Maduwa and on 22^nd^ and 23^rd^ November 2024 in Baragu during the short rainy season. Presence of anopheline and culicine larvae was recorded as early (1^st^ and 2^nd^) instars or late (3^rd^ and 4^th^) instar larvae on a field data form. Presence of mosquito pupae in habitats was also recorded. Aquatic habitats were classified based on their appearance as either natural swamp, cultivated swamp, river fringe, puddle, open drain or burrow pits. Perimeter of water surface in each habitat was estimated and recorded either as <10m, 10-100m or >100m while water depth was measured using a meter rule and recorded either as <50cm or >50cm. Coordinate readings (latitude and longitude) and altitude of aquatic habitats were taken using the GPS unit.

### Adult mosquito sampling

Indoor resting mosquitoes were sampled using pyrethrum spray catches (PSC) method in ten randomly selected houses once per site following standard WHO protocols [27]. Sampling was conducted on 8^th^ November 2024 in Maduwa and on 25^th^ November 2024 in Baragu during the short rainy season. This was conducted in all rooms of the ten randomly selected houses after obtaining consent from either the household-head or his/her adult representative. Household characteristics, including number of occupants the previous night, use of insecticide-treated nets or coils, presence of ceilings, and whether eaves were closed, were recorded.

### Mosquito identification

Collected adult mosquitoes were identified into their species using morphological features [28,29] in the field, counted and recorded on a field data form. Female mosquitoes were recorded based on their gonotrophic stages either as unfed, blood-fed, half-gravid or gravid. Adult *Anopheles* mosquitoes were preserved in silica gel and transported to KEMRI-CGHR laboratories. Due to unavailability of funds, molecular identification of these *Anopheles* mosquitoes to sibling species was not done.

### Study participants

Human participants, both male and female, were eligible to be enrolled in this study. Inclusion/exclusion criteria included: resident within the selected study area; aged one year and above for both Maduwa and Baragu; aged 18 years and over in Budalangi; provided signed informed consent for adults and assent for children aged 7 – 17 years, willingness to provide information about gender, age and weight measured and then a blood sample taken by finger-prick method. Budalangi, a second site in Busia County, was included to screen adult human participants only for malaria parasites with the aim of identifying gametocyte donors for another separate objective of this study.

### Malaria blood-smear sampling and testing

Sampling for malaria blood-smears was conducted on 7^th^ and 8^th^ November 2024 in Maduwa and from 14^th^ November 2024 to 7^th^ December 2024 in Baragu during the short rainy season. A household-head or his/her adult representative was identified, consented and interviewed to provide information about his/her family members aged one year and above using a questionnaire form in Open Data Kit (ODK). This information included the number of family members, their gender and age. After this, each adult member of the family was individually consented and children aged 7-17 years assented. A one-time finger-prick was done by a certified phlebotomist to collect malaria thick and thin blood smears for microscopic examination and tested on site using a malaria rapid diagnostic test (mRDT) kit. Malaria screening among adults only in Budalangi and its environs was conducted from 16^th^ January 2025 to 20^th^ January 2025 and from 25^th^ March 2025 to 28^th^ March 2025. Malaria screening camps were stationed in each of the nine surrounding villages, and engaged local Village Elders and Community Health Promoters (CHPs) to invite adult volunteers to participate. Participants who tested malaria positive with uncomplicated malaria were given artemether-lumefantrine and paracetamol by a local CHP according to the current Kenyan Ministry of Health recommended case management guidelines [30].

### Microscope examination

Malaria blood-smear slides from Maduwa and Baragu were transported to KEMRI-CGHR laboratories and the slides from Budalangi and its surrounding villages were taken to Budalangi Health Centre where they were stained using 10% Giemsa stain. Malaria microscopy by expert microscopists was performed using Giemsa-stained thick and thin blood smears, examined under a light microscope at ×1000 magnification (oil immersion) to detect and identify *Plasmodium* species as *Plasmodium falciparum*, *P. malariae* or *P. ovale*. Malaria parasites/gametocytes life-cycle stage of parasites were counted against 200 white blood cells (WBCs) and converted to the number of malaria parasites per microlitre (MPS/µl) of blood by using the formula (Number of parasites / Number of white blood cells counted (200)) x 8,000, using the assumption of 8,000 white blood cells (WBCs) per µl of blood [31].

### Rainfall, temperature and relative humidity

A rain gauge, air temperature and relative humidity data loggers (HOBO^®^ Onset data loggers, Onset Computer Corporation 470 Bourne, MA, USA) were used to collect climate data from 1^st^ November 2024 to 30^th^ April 2025. The rain gauge was installed on an iron stand one and a half metre off the ground within the 1km x 1km study area in Maduwa and Baragu and it collected data as per the rainfall events. A temperature/relative humidity data logger was hanged on a string within the field insectary with netted meshed walls (used for another objective of this study) to record ambient temperature and relative humidity at one-hour intervals. Following failure of the temperature/relative humidity logger at the Baragu site in January 2025, supplementary daily temperature and relative humidity data for February, March and April 2025 were obtained from the Kenya Meteorological Department station at Kenya Methodist University (KeMU) in Meru Town. Due to this, data collected from 1^st^ November 2024 to 9^th^ January 2025 was used to calculate daily minimum, average and maximum temperatures and relative humidity.

### Ethical considerations

Ethical approval for this research study was obtained from the Kenya Medical Research Institute – Scientific and Ethical Review Unit (KEMRI-SERU) (KEMRI/SERU/CGHR/01-07-470/4804). Research license was obtained from the National Commission for Science, Technology & Innovation (NACOSTI) (License No: NACOSTI/P/24/35942). Approval to conduct this research study was obtained from the County Commissioners in Busia (Ref. No. ADM 15/27 VOL.I/159) and Meru (Ref. EDU.12/3 VOL IV (143)) counties. These administrative approval letters were submitted to Deputy County Commissioners, Assistant County Commissioners, Chiefs and Assistant Chiefs in Maduwa, Baragu and Budalangi study sites. This research study was introduced to local residents through public meetings (*barazas*) in each of the study sites and villages. Individual informed consent and assent, where applicable, were obtained before any information and/or blood samples were collected from any study participants, their houses and parcels of land.

### Data management and analysis

All data were collected on field and laboratory forms (on paper or in ODK), entered in Microsoft Excel databases, cleaned and then backed up in Dropbox. The difference in the presence of *Anopheles* larvae in aquatic habitats and the number of people who tested malaria positive using the mRDTs in Maduwa and Baragu were analysed using Chi-Square (χ^2^). Logistic regression analysis was used to determine the likelihood of finding *Anopheles* larvae in aquatic habitats in these sites. The difference in the number of adult mosquitoes in houses, the number of malaria parasites per microlitre (MPS/µl) of blood and rainfall, temperature and relative humidity data among the study sites were analysed using analysis of variance (ANOVA). Raw temperature data for Baragu and Maduwa were sorted ascending and all the hours in which temperatures recorded were below 16°C, the annual mean minimum temperature required for *P. falciparum* transmission [16,17,19], were selected, grouped in count of continuous hours (1 hour - 10 hours, the maximum count that was recorded) and then their frequencies were counted. Analyses were performed in Statistical Package for the Social Sciences (SPSS) Version 22 and Stata Version 19.

## Results

### Study population

A total of 751 participants were enrolled in the study: 80 (10.7%) from Maduwa, 284 (37.8%) from Baragu, and 387 (51.5%) from Budalangi and its surrounding villages. Overall, 431 (57.4%) participants were female and 320 (42.6%) were male. In Maduwa and Baragu, participants were aged ≥1 year, whereas in Budalangi and neighbouring villages, eligibility was restricted to adults aged ≥18 years (Tables 1 and 2). In Maduwa, 38 (47.5%) participants were female and 42 (52.5%) were male, with a mean age of 34.3 years (34.1 years for females and 34.5 years for males). In Baragu, 151 (53.2%) were female and 133 (46.8%) were male, with an overall mean age of 49.4 years (53.6 years for females and 44.6 years for males). In Budalangi and its environs, 242 (62.5%) were female and 145 (37.5%) were male, with a mean age of 49.0 years (49.2 years for females and 48.8 years for males). In Maduwa and Baragu, body weights ranged from 8.1 kg to 119.6 kg, with a mean weight of 53.0 kg. In Budalangi and surrounding villages, weights ranged from 34.4 kg to 130.6 kg, with a mean of 65.4 kg. A total of 381 residential houses were mapped within the 1 km × 1 km selected study areas across the two sites: 330 (85.3%) in Baragu and 51 (13.2%) in Maduwa. In Baragu, 299 (90.6%) houses were occupied, 26 (7.9%) were unoccupied, and 5 (1.5%) were abandoned. In Maduwa, 35 (68.6%) houses were occupied, while 16 (31.4%) were abandoned, largely due to perennial flooding.

**Table 1.**
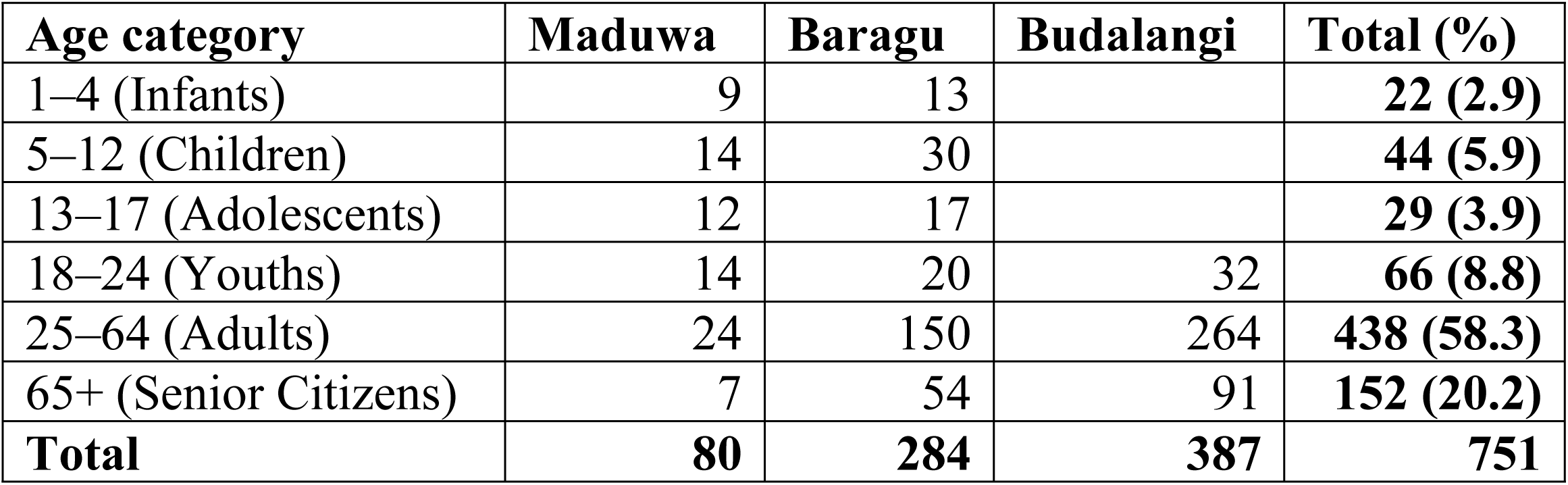
Total number of study participants and their age categories.

| Age category | Maduwa | Baragu | Budalangi | Total (%) |
| --- | --- | --- | --- | --- |
| 1–4 (Infants) | 9 | 13 |  | **22 (2.9)** |
| 5–12 (Children) | 14 | 30 |  | **44 (5.9)** |
| 13–17 (Adolescents) | 12 | 17 |  | **29 (3.9)** |
| 18–24 (Youths) | 14 | 20 | 32 | **66 (8.8)** |
| 25–64 (Adults) | 24 | 150 | 264 | **438 (58.3)** |
| 65+ (Senior Citizens) | 7 | 54 | 91 | **152 (20.2)** |
| **Total** | **80** | **284** | **387** | **751** |

**Table 2.** Total number of study participants in Budalangi and its neighbouring villages.

| Number | Villages in Budalangi area | Total (%) |
| --- | --- | --- |
| 1 | Budunyi | 77 (19.9) |
| 2 | Bulagu | 35 (9.0) |
| 3 | Gimini | 49 (12.7) |
| 4 | Ikhula | 37 (9.6) |
| 5 | Khulwano | 33 (8.5) |
| 6 | Mubwayo | 73 (18.9) |
| 7 | Mulwa | 40 (10.3) |
| 8 | Other villages* | 43 (11.1) |
|  | **Total** | **387** |

\*Other villages = Buchiriba (5), Bujonjoli (7), Bugisa (1), Bukholi (1), Busidia (3), Busweti (8), Mang'ong'o (2), Mukhobola (7), Mumbira (1), Nagoba (3), Ruambwa (1) and Sikokhe (4).

Ground truthing of the study sites indicated that the landscape in Baragu is mainly undulating steep hills and mostly narrow “V-Shaped” valleys. Maduwa is at the shore of Lake Victoria and is mainly flat wetland marshy plains characterized by saturated soils, slow-moving and standing water dominated by Napier grass and papyrus reeds. Budalangi is also close to Lake Victoria and is defined by low-lying, flat landscape within the floodplains of the River Nzoia.

### Larval stages

A total of 99 aquatic habitats were identified: 32 (32.3%) in Baragu and 67 (67.7%) in Maduwa (Fig 1). Most (82.8%) of these aquatic habitats were cultivated swamps, drains and pools (Table 3). Vegetation cover in these aquatic habitats ranged from 0% (9.1%), 1-25% (16.2%), 26-50% (15.2%), 51-75% (22.2%) to 76-100% (37.4%). Over a half (53.5%) of these aquatic habitats were small in size (<10m). Habitats 10-100m in perimeter were 29.3% while the largest ones (>100m) were 17.2% and were found only in Maduwa. Almost all (90.9%) of the aquatic habitats were shallow (<50cm deep). In both Baragu and Maduwa, *Anopheles* larvae were found breeding in a third (29.3%) of all the aquatic habitats and were present mostly (82.8%) in swamps and drains. In Baragu alone, they were found breeding in swamps, puddles, pools and drains. None was found breeding in cemented pits. In Maduwa alone, they were found breeding in all types of aquatic habitats, namely, swamps, river-fringes, puddles, pools, burrow-pits and drains. The presence of *Anopheles* early instar larvae in aquatic habitats in Maduwa and Baragu was significantly different (χ^2^ (df=1, N = 99) = 4.26, p = 0.04). However, the presence of *Anopheles* late instar larvae in these habitats in the two sites was not significantly different (χ^2^ (df=1, N=99) =2.43, p=0.12). Other variables, namely, habitat type, vegetation cover, water depth and size of the water body were not significantly associated with the presence of *Anopheles* larvae in the aquatic habitats. Logistic regression results revealed that Baragu (15.6%, n=5/32) was less likely to have *Anopheles* early instar larvae in aquatic habitats compared to Maduwa (35.8% n=24/67; Unadjusted Odds Ratio (uOR)=0.33; 95%CI, 0.11-0.97; P=0.05).

**Table 3.** Type and number of aquatic habitats in Baragu and Maduwa sites.

| Habitat type | Baragu | Maduwa | Total (%) |
| --- | --- | --- | --- |
| Cultivated swamps | 6 | 31 | **37 (37.4)** |
| River fringes | 2 | 2 | **4 (4.0)** |
| Puddles | 3 | 3 | **6 (6.1)** |
| Pools | 9 | 5 | **14 (14.1)** |
| Borrow pits | 1 | 5 | **6 (6.1)** |
| Drains | 10 | 21 | **31 (31.3)** |
| Cemented pits | 1 | 0 | **1 (1.0)** |
| **Total** | **32** | **67** | **99** |

### Adult mosquitoes

A total of 58 individuals slept in the houses the previous night before indoor resting mosquito sampling using PSC method: 22 (37.9%) were in Baragu and 36 (62.1%) in Maduwa. Most of them were adults (31, 53.4%) than children (27, 46.6%). On average, more people slept in the houses in Maduwa (3.6) than in Baragu (2.2). No insecticide was sprayed in the houses nor mosquito coil used. Out of the 10 houses sampled per site, most of occupants in Maduwa (7) slept under a treated bed net than in Baragu (3). Half (5) of the houses in Baragu had closed bedroom eaves and two in Maduwa. Two houses in Baragu and also two in Maduwa had a ceiling in their bedrooms. Spatial repellents, provided by another research study, were used in 9 out of the 10 houses only in Maduwa.

A total of 96 adult mosquitoes were collected by PSC sampling method: 21 (21.9%) in Baragu and 75 (78.1%) in Maduwa. Out of these mosquitoes, 4 (4.2%) were *An. gambiae*; 2 (2.1%) *An. funestus*; 82 (85.4%) *Culex* and 8 (8.3%) *Mansonia uniformis*. The four *An. gambiae* mosquitoes were 3 males and 1 blood-fed female. One male was collected in Baragu and two in Maduwa as well as the single female mosquito. The two *An. funestus* mosquitoes were collected in Maduwa only: one male and one blood-fed female. The 82 *Culex* mosquitoes were collected in Baragu (20, 24.4%) and in Maduwa (62, 75.6%). They were: 25 (30%) males; 33 (40%) unfed; 14 (17%) blood-fed; 6 (7%) half-gravid and 4 (5%) gravid. In Baragu, 10 (50%) of them were females whereas they were 47 (76%) in Maduwa. Eight female *Mansonia uniformis* mosquitoes were collected in Maduwa only: six were unfed whereas two were half-gravid. There was no statistical difference in combined mean densities of malaria vectors (*An. gambiae* and *An. funestus*); between Maduwa and Baragu sites (t(18)=1.3805, p=0.18). Adult mosquito collections were low, limiting statistical power to detect differences in indoor resting densities.

### Malaria RDT

A total of 651 participants were screened using mRDT, of whom 37 (5.7%) tested positive for malaria. The prevalence varied by site: 0% (0/250) in Baragu, 12.5% (9/72) in Maduwa, and 8.5% (28/329) in Budalangi and its surrounding villages. This difference was statistically significant (χ² (df = 2, N = 651) = 26.211, p<0.001).

### Malaria microscopy

A total of 69/709 (9.7%) of the participants tested positive for malaria based on malaria microscopy. Malaria prevalence by microscopy was 2.8% (2/71) in Maduwa and 17.3% (67/387) in Budalangi and its surrounding villages. The malaria prevalence in Budalangi in January 2025 was 17.1% (36/210) whereas it was 17.5% (31/177) in March 2025. The average and maximum number of malaria parasites per microlitre (MPS/µl) of blood was 258.5 and 8400 in Budalangi and 148.6 and 5040 in Maduwa. The differences in the number of malaria parasites per microlitre were statistically significant among the three study sites (F(2,706)= 9.477, p<0.001). No participant tested positive (0/251) by microscopy in Baragu.

### Rainfall

Within the six months (November 2024 to April 2025), Maduwa received more rainfall (725.9 mm) than in Baragu (428.6 mm) (Fig 2). The total monthly rainfall that was received in Maduwa was 300.0 mm in November 2024, 106.9 mm in December 2024, 17.5 mm in January 2025, 0.3 mm in February 2025, 79.5 mm in March 2025 and 221.7 mm in April 2025. Similarly, the total monthly rainfall that was received in Baragu was 44.5 mm in November 2024, 3.6 mm in December 2024, 0.3 mm in January 2025, 13.8 mm in February 2025, 213.6 mm in March 2025 and 152.9 mm in April 2025. Average daily rainfall in Maduwa was 4.0 mm (95% Confidence Interval 2.5 - 5.5) and in Baragu it was 2.4 mm (1.2 - 3.5), which were not statistically different (F(1,360)= 2.899, p=0.09).

**Fig 2.**
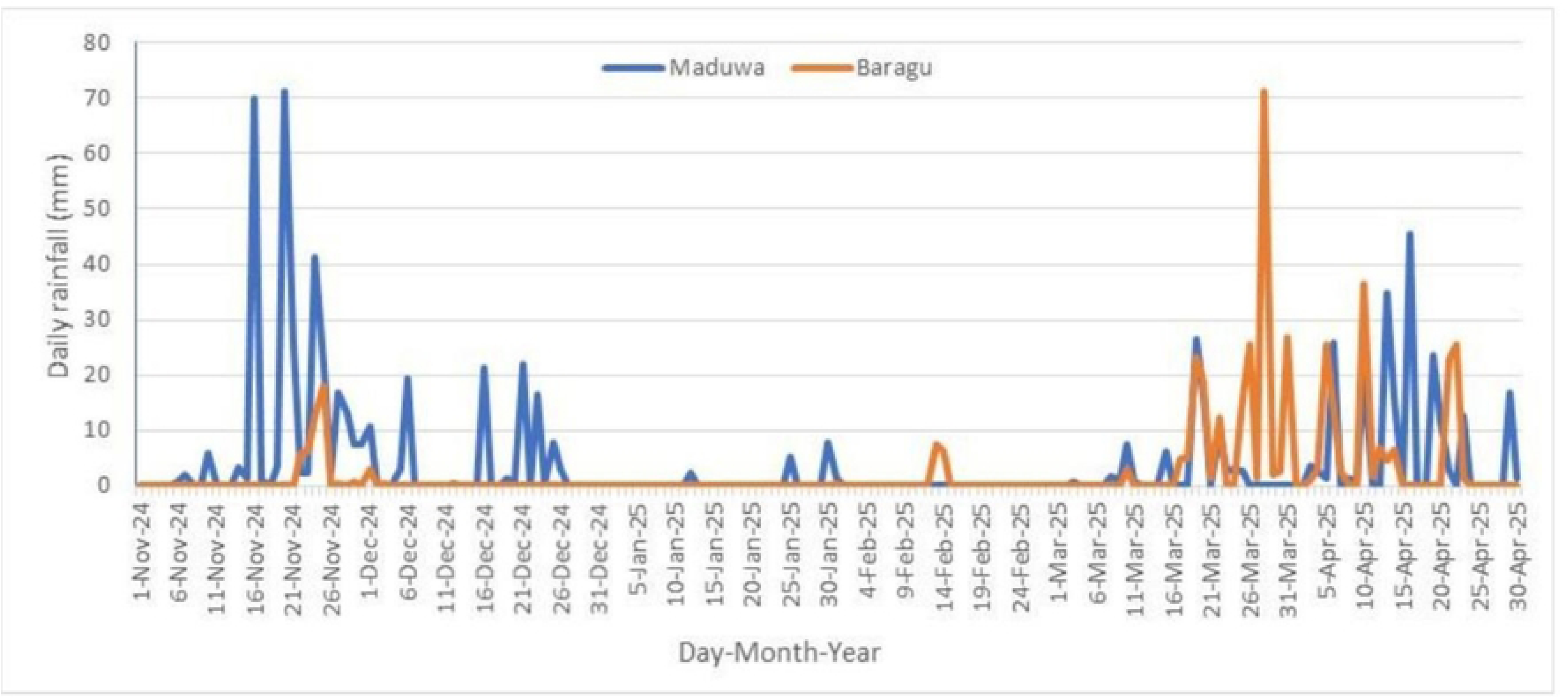
Daily rainfall received in Maduwa and Baragu study sites.

### Temperature

Minimum, average and maximum temperatures in Baragu were 10.0°C, 20.7°C and 36.5°C, respectively (Table 4). Similarly, in Maduwa they were 13.7°C, 23.4°C and 35.1°C, respectively. On almost all the days, temperatures were higher in Maduwa than in Baragu (Fig 3). Daily minimum temperatures in Maduwa were higher than in Baragu with an average difference of 2.6°C (19.2°C - 16.6°C) (Fig 4). Similarly, daily average temperatures in Maduwa were higher than in Baragu with an average difference of 1.9°C (23.3°C - 21.4°C) (Fig 4). Interestingly, daily maximum temperatures in Maduwa and Baragu were the same during the night, morning and evening hours, but they were higher in Baragu than in Maduwa during the day between 8:00AM to 3:00PM with an average difference of 1.2°C (30.4°C - 29.2°C) (Fig 4). The average minimum, mean and maximum temperatures were significantly higher in Maduwa than in Baragu, F(1,360)= 89.505, p<0.001, F(1,360)=187.741, p<0.001 and F(1,360)=161.706, p<0.001, respectively. Despite the fact that temperature data in Baragu was recorded from November 1^st^ 2024 to January 9^th^ 2025 (2 months 9 days), due to the data logger failure, as opposed to Maduwa where it was recorded successfully from November 1^st^ 2024 to April 30^th^ 2025 (6 months), Baragu had significantly (F(1,18)= 9.656, p=0.01) more hours below 16°C than Maduwa (Table 5).

**Fig 3.**
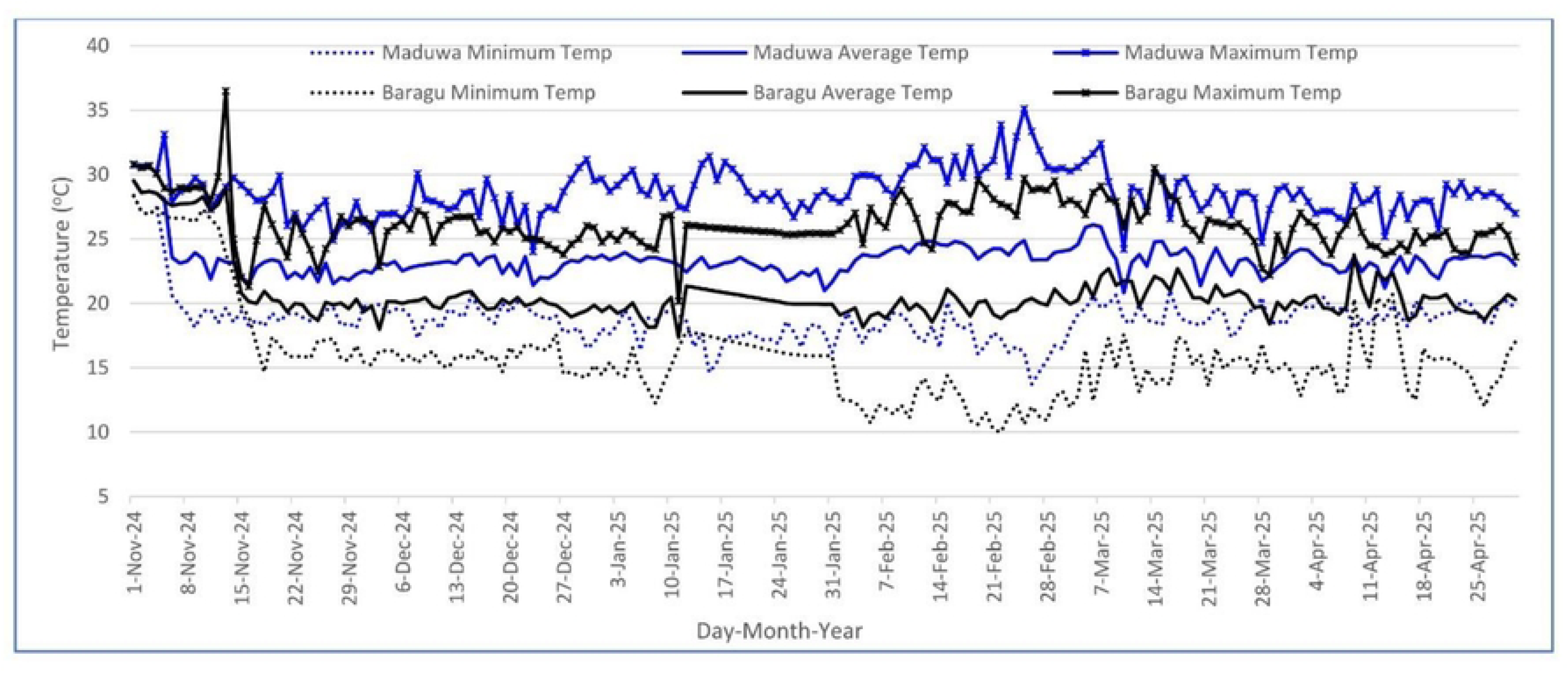
Daily minimum, average and maximum temperatures recorded in Maduwa and Baragu study sites.

**Fig 4.**
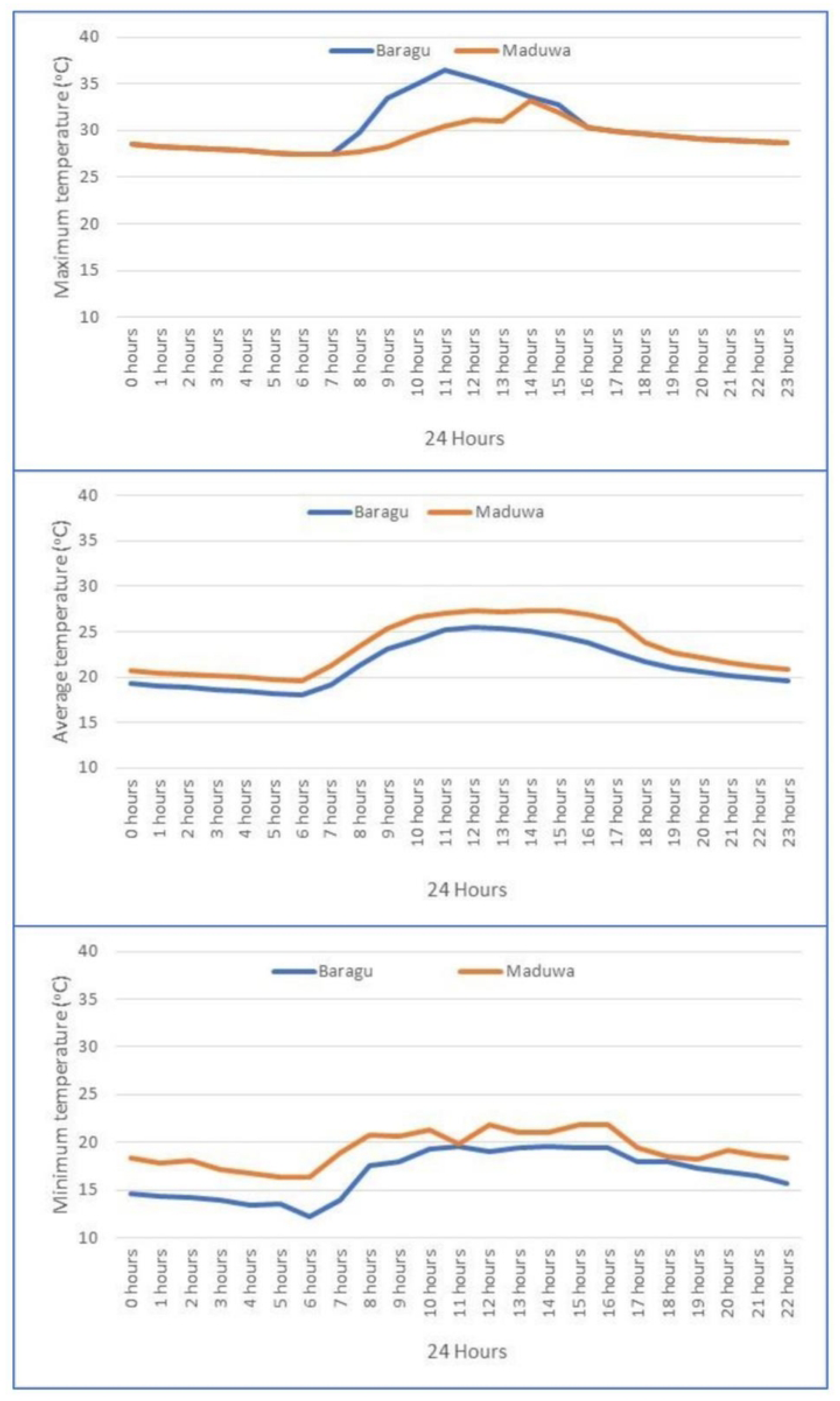
24-hour minimum, average and maximum temperatures recorded in Maduwa and Baragu study sites.

**Table 4.** Minimum, average and maximum temperatures recorded in Baragu and Maduwa sites.

| Site | Summary | Minimum Temperature °C | Average Temperature °C | Maximum Temperature °C |
| --- | --- | --- | --- | --- |
| Baragu | Minimum | **10.0** | 17.4 | 20.2 |
|  | Average | 15.9 | **20.7** | 26.2 |
|  | Maximum | 27.5 | 29.0 | **36.5** |
| Maduwa | Minimum | **13.7** | 20.8 | 24.0 |
|  | Average | 18.8 | **23.4** | 28.7 |
|  | Maximum | 27.5 | 28.7 | **35.1** |

**Table 5.** Frequency of continuous hours below 16°C ambient temperature in Baragu and Maduwa sites.

| Frequency of hours below 16°C | Baragu |  | Maduwa |  |
| --- | --- | --- | --- | --- |
|  | Count hours | Total hours | Count hours | Total hours |
| 1 | 8 | 8 | 2 | 2 |
| 2 | 14 | 28 | 2 | 4 |
| 3 | 21 | 63 | 6 | 18 |
| 4 | 24 | 96 | 4 | 16 |
| 5 | 5 | 25 | 0 | 0 |
| 6 | 18 | 108 | 0 | 0 |
| 7 | 0 | 0 | 0 | 0 |
| 8 | 0 | 0 | 0 | 0 |
| 9 | 18 | 162 | 0 | 0 |
| 10 | 10 | 100 | 0 | 0 |
| **Total** | **118** | **590** | **14** | **40** |

\*Note: Data from Baragu was collected from November 1<sup>st</sup> 2024 to January 9<sup>th</sup> 2025 (2 months and 9 days) due to hourly temperature data logger break down, while data from Maduwa was collected from November 1<sup>st</sup> 2024 to April 30<sup>th</sup> 2025 (6 months, the entire study period).

### Relative humidity

Minimum, average and maximum levels of relative humidity in Maduwa were 21.6%, 78.7% and 99.3%, respectively. Similarly, they were 30.3%, 73.3% and 96.9%, respectively, in Baragu. Daily minimum, average and maximum levels of relative humidity in Maduwa were higher than in Baragu with an average difference of 4.1% (46.7% - 42.6%), 5.4% (79.2% - 73.8%) and 2.3% (94.9% - 92.6%), respectively (Fig 5). The average minimum RH in Baragu was significantly higher than in Maduwa (F(1,360) =19.376, p<0.001). Both average mean and maximum RH levels were significantly higher in Maduwa than they were in Baragu, (F(1,360)=87.687, p<0.001) and (F(1,360)=240.441, p<0.001), respectively.

**Fig 5.**
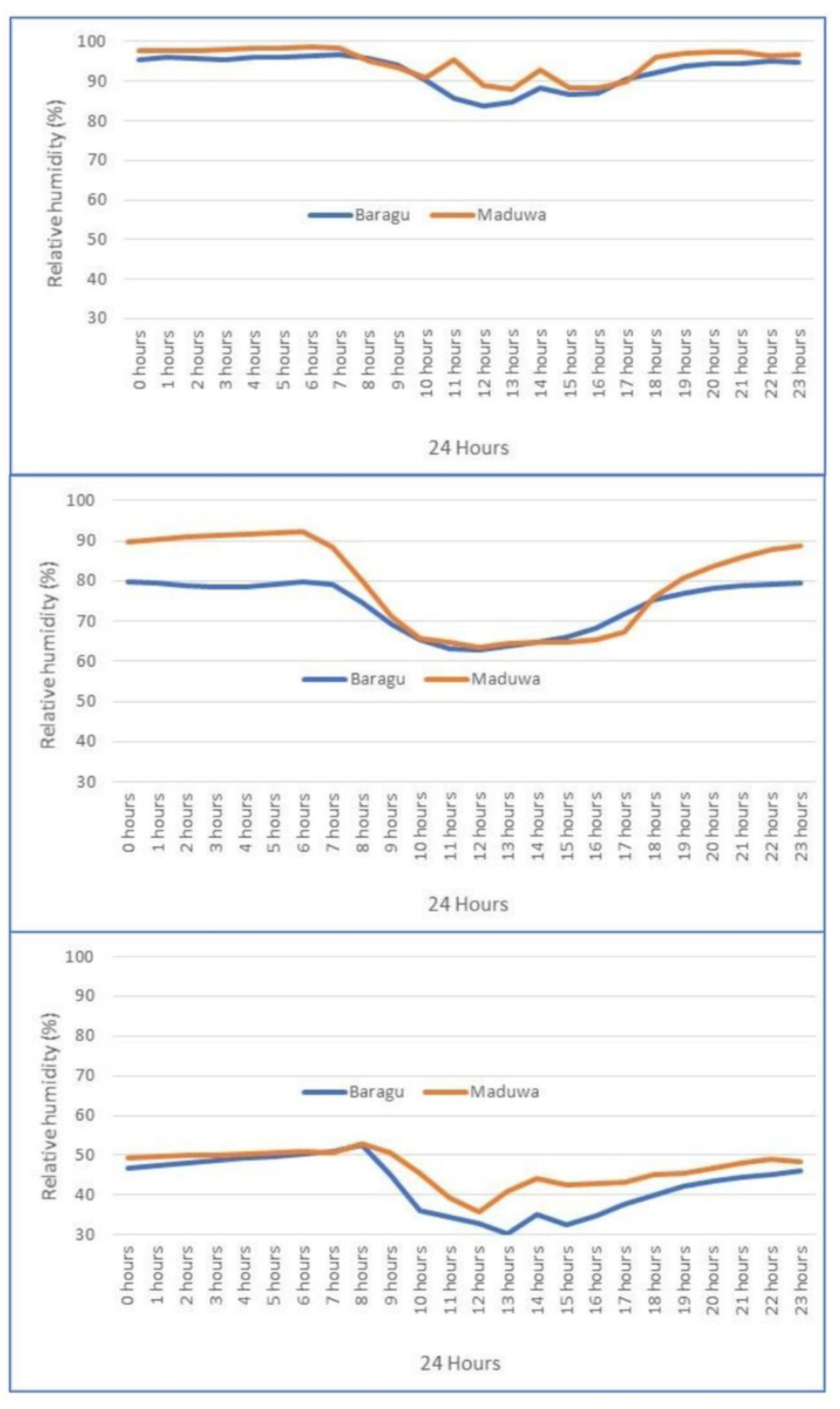
24-hour relative humidity recorded in Maduwa and Baragu study sites.

## Discussion

The first finding of this research study is the continued absence of malaria cases in Baragu at an altitude that ranged from 1,411-1,531 m in Meru County within the central Kenya highlands. This finding is similar to the report by Gichuki et al [32] who did not detect any malaria case in Mwea within this region. This confirms that there are areas within the central Kenya highlands that are still correctly characterized as malaria-free [13,14] and forms most part of the central Kenya highland region. It is important to note that we found few *An. gambiae s.l.*, both in larval and adult stages, breeding in Baragu. In 2005, a study by Chen et al [33] identified *An. arabiensis* both in Karatina and Naro Moru within this central Kenya highlands at elevations of 1,720-1,921 m. This highlights the existing potential risk of local malaria transmission should malaria parasites be introduced in these areas. Recently, Kimani et al [34] identified malaria vectors (*An. funestus* group, *An. ziemenni*, *An. gambiae*, *An. christyi*, *An. coustani* and *An. demeilloni*) breeding in Karai Rurii and Ondiri Swamp in Kikuyu at higher altitude that ranged from 1,980-1,998 m in the central Kenya highlands, west of Nairobi. In addition, they reported local malaria transmission at a prevalence of 5.6% (47/838). This indicates that local malaria transmission is making entries at higher altitudes in parts of the central Kenya highlands. Malaria vectors have established in areas surrounding Mt. Kenya at an altitude ranging from 1,100-2,000 m within the central Kenya highlands making the population at risk of local malaria transmission. Finding malaria cases both in Maduwa and in Budalangi and its environs in Busia County was expected as they are located within the western Kenya malaria endemic zone that has the highest prevalence and risk of infection in the county [9–12,35].

The second finding of this research study is that Baragu recorded a total of 590 hours in minimum temperature below 16°C, in slightly more than a third of the time period in which Maduwa recorded 40 hours. This temperature has been associated with inhibition of sporogonic development of malaria parasites in *Anopheles* mosquitoes in this area. While it has been documented that the annual mean minimum temperature required for *P. falciparum* transmission is 16°C [16,17,19], it would be necessary to establish the actual number of hours in which the minimum temperature experienced is below this mean as we attempted to do in this study. The use of station-based temperature data for Baragu after logger failure may have reduced microclimatic resolution and potentially underestimated local temperature variability. Therefore, more work is needed to generate annual hourly temperature data to support this view. Ongoing climate change is expected to cause increases in temperature within high altitude areas like the central Kenya highlands [36]. This may create conducive temperatures for malaria vectors to successfully adapt, survive and thrive at higher altitudes in this area. This may ultimately lead to an increase in the prevalence and the geographical distribution of mosquito-borne diseases like malaria in the Kenyan highlands. The average relative humidity recorded both in Maduwa (78.7%) and in Baragu (73.3%) were within the required range of 60-100% for the highest survival of adult *An. gambiae s.s.* mosquitoes [18]. A study by Omukunda et al [37] reported an association between the abundance of *An. gambiae s.s.* and relative humidity of both the same month and the previous month in the highland site in western Kenya but not in the low land site. The total amount of rainfall that was received in Maduwa was more than the amount that was received in Baragu. Considering rainfall alone, one would expect to have more malaria cases in Maduwa than Baragu. This is because, there is a positive association between rainfall and malaria cases within the lowlands surrounding Lake Victoria but not in the adjacent highland areas [38].

The third finding is that topography is one feature that contributes to define levels of malaria transmission in an area. In Baragu, valleys are generally narrow (V-shaped) with numerous streams. In addition to the V-shaped valleys in the highlands, some areas are characterized by broad flat-bottomed (U-shaped) valleys and plateaus [20,22]. In the V-shaped valleys of Baragu, we recorded few mosquito breeding habitats, presence of *Anopheles* larvae in few habitats, few *Anopheles* adult and no malaria infection. This is consistent with the reports of fewer number of *Anopheles* larvae/dip in habitats, fewer adult vector densities/house, fewer malaria prevalence, fewer distribution of infections and people less immune to malaria among residents of V-shaped valleys compared to U-shaped ones within the western Kenya highlands [20–22]. Maduwa and Budalangi are located in generally flat areas in the lower courses of River Yala and River Nzioa, respectively, that are prone to flooding during the rainy seasons [39]. Floods have been reported to cause an increase both in malaria cases and mortality [24,40,41]. They create more fresh aquatic habitats especially in the upper course of rivers by saturating the soils, enhancing the breeding of more malaria vectors that initiate and sustain increased local malaria transmission [24]. However, in the lower course of a river, floods are known to initially wash away mosquito aquatic stages, a situation known as the “flushing effect”, hence reducing malaria cases [23]. After a lagged period of weeks, a month or two when flood waters subside forming pools, malaria vectors re-establish themselves and lead to increased malaria transmission [24,42]. This might be the reason we recorded few malaria cases in Maduwa that experiences more flooding compared to Budalangi.

The fourth finding is the malaria prevalence of 17.3% (67/387) among adult residents in Budalangi and its surrounding villages in January and March of 2025 during the dry season and towards the onset of the main rainy season. This finding confirms that malaria transmission persists at a considerable level throughout the dry season when breeding habitats are greatly reduced. Amare et al [43] reported comparable malaria prevalence of 16.4% during the dry season in Ethiopia. Collins et al [44] demonstrated that school aged children and adults who have likely experienced repeated malaria infections form the main reservoir for malaria transmission during the dry season. In another study, Jawara et al [45] reported the presence of malaria vectors throughout the dry seasons but in low numbers. During the dry season, most of these mosquitoes breed in pools that are formed by drying streams [44]. The presence of these vectors during dry periods plays an important role in sustaining malaria transmission until the onset of the rainy season.

The first limitation of this research study was that we sampled malaria cases and vectors during the short-rainy season in November and December in 2024 and end of the dry season in January and March 2026, hence we recorded low malaria prevalence and collected few vectors. The initial plan was to start sampling on 1^st^ March and end on 31^st^ July but this was delayed by planning preparations. Therefore, we missed out on the main malaria transmission season that normally occurs from April to July [25,46]. The second limitation is that our temperature data logger in Baragu broke down, hence we missed to collect hourly data according to plan. The alternative data we purchased from the Kenya Meteorological Department was only one entry per day. This did not enable us to appropriately compare the two data sets.

## Conclusion

No malaria cases were reported in Baragu in Meru County and few cases were reported both in Maduwa and Budalangi in Busia County, which persist throughout the dry season. Although *Anopheles* mosquitoes were found breeding in aquatic habitats, few adult mosquitoes were collected in Maduwa and Baragu. More hours below 16°C and generally narrow (V-shaped) valleys in Baragu and generally flat areas in the lower courses of rivers that are prone to flooding during the rainy seasons in Maduwa and Budalangi, partly explain for the low malaria cases reported in this study. Future longitudinal studies spanning peak transmission seasons are needed to validate these patterns across years.

## Data Availability

All data produced in the present study are available upon reasonable request to the authors.

## Acknowledgements

We thank the local residents and administrators in the study sites in Baragu in Meru County and Maduwa and Budalangi in Busia County for facilitating the necessary approvals to conduct all field activities of this research study. We particularly thank all local study participants who volunteered and offered their assistance during all data collection activities. We appreciate all assistance offered by Kenya Medical Research Institute staff and all project staff in Baragu, Maduwa and Budalangi. This paper is published with the permission from the Director General Kenya Medical Research Institute.

## Author contributions

Conceptualisation: BAN, GAA, ED, LWK, HMA; Methodology: BAN, GAA, KOO, JOM, HMA; Writing – original draft: BAN, GAA, KOO; Writing – review & editing: BAN, GAA, KOO, JOM, ED, LWK, HMA; Project administration: BAN; Funding acquisition: BAN, GAA, ED, LWK, HMA.

